# Social and Societal Factors Interact with Psychological Factors to Shape Pain Outcomes in a Community Sample with Chronic Pain: A Network Study

**DOI:** 10.1101/2025.08.29.25334446

**Authors:** Gwen van der Wijk, Lisette de Groot, Joline Bruweleit, Lilla Hévizi, Meredi S. Edgarian, Stijn B. Vermeulen, Lakeya S. McGill

## Abstract

Although the biopsychosocial model of chronic pain is widely recognized, few studies include social-societal factors. Here, we examined how these interact with psychological factors to shape pain outcomes. We collected self-reported data from 262 participants with chronic pain about emotional states, pain coping strategies, social interactions, societal stressors and pain outcomes. After replicating previous partial correlation networks including only psychological factors, we extended our analysis to include social and societal factors, which demonstrated that these had strong direct relationships with quality of life and were indirectly related to pain intensity and pain disability. Our results indicate that social and societal factors are important for understanding pain outcomes and should be considered in interventions targeting chronic pain. Future work should examine the interactions among social-societal and psychological factors in more depth to inform ways to incorporate this into individual pain management and societal interventions for chronic pain.

## Introduction

Chronic pain, defined as persistent or recurrent pain for three months or more (Treede et al., 2015), is a disabling condition affecting individuals all over the world. The overall prevalence of chronic pain in Europe is approximately 20%, and studies show an increase in its global prevalence, with higher numbers expected for certain subtypes in the regions South Asia, Tropical Latin America and North America in the future (Rometsch et al., 2024; Zhu et al., 2025). The prevalence of chronic pain also differs between socio-demographic groups within countries. For example, a systematic review of European studies found a higher prevalence of chronic pain among women, individuals with a low income, poor social support, and Asian and Black people, stressing the importance of taking sociodemographic dimensions into account (Rometsch et al., 2025).

Although sociodemographic variables are associated with chronic pain, they do not inherently explain the underlying causes of differences in prevalence. The World Health Organization (WHO) addresses this with its notion of ‘social determinants,’ in which they explain that health inequalities arise “*[…] because of the circumstances in which people grow, live, and age, and the systems put in place to deal with illness. The conditions in which people live and die are, in turn, shaped by political, social, and economic forces*” (World Health Organization, 2008, p. 3). In this article, we use ‘social indicators’ to reflect that these circumstances, while influential, do not predetermine any individual’s outcomes but can change and be changed (Salerno and Bogard, 2019). In line with this perspective, we look at specific social and societal experiences instead of demographic group membership, as this may reveal more direct and useful insights (e.g., looking at experiences of discrimination instead of racial identity; Letzen et al., 2022).

In the majority of psychological research on chronic pain, such social indicators are not taken into account (Kapos et al., 2024). Instead, the focus tends to be on individual psychological characteristics. For example, a popular theory of chronic pain is the fear-avoidance model, which states that pain-related worrying, as well as the subsequent pain-related fear and avoidance behaviors, can lead to higher pain disability and worse pain (Vlaeyen and Linton, 2012). Many studies have supported this model (e.g., Rogers and Farris, 2022), and it has led to the development and implementation of exposure therapy for chronic pain (Vlaeyen et al., 2012). Higher levels of anxious and depressive symptoms have also been observed in people experiencing chronic pain, and this has been associated with worse pain outcomes (Burke et al., 2015).

The importance of positive psychological factors in chronic pain is also increasingly recognized (Blasco-Belled et al., 2025). Broadly, the idea is that factors such as positive affect can help people manage their chronic pain and improve their quality of life. A prominent theory is the psychological flexibility model, which focuses on adjusting responses to thoughts and feelings associated with pain instead of trying to change the pain-related thoughts and feelings themselves (McCracken and Morley, 2014). For example, it proposes that higher pain acceptance will allow someone to move from fighting the pain to investing their energy into doing valued activities despite the pain. This model has been translated into acceptance and commitment therapy (ACT) for chronic pain (Scott and McCracken, 2015).

Although many studies show associations between these psychological factors and important pain outcomes, and psychological interventions targeting these factors have been found effective, treatment effects are generally small to moderate and vary between individuals (McCracken, 2023; Williams et al., 2020). One potential reason for these limited results may be that the frameworks such treatments are based on largely ignore the social-societal context (Kapos et al., 2024). Indeed, foundational and recent work from both a sociology and critical medical anthropology perspective indicates that social, cultural and political context is essential for understanding health, as it reflects, is shaped by and deeply embedded in these contexts (Gooberman-Hill, 2015; Kleinman, 1992; Scheper-Hughes and Lock, 1987; Webster et al., 2023; Zajacova et al., 2021). Bringing this context into the picture may bring a better understanding of the roots of variation in psychological factors, and through that, may reveal ways to make therapy more nuanced and sensitive to the social, cultural and political context individuals navigate (Ashton-James et al., 2022; Kapos et al., 2024).

Partial correlation networks examine the unique relationships between pairs of variables while controlling for all others in the network, and are therefore well suited to explore the complex interactions and relative contributions of multiple factors. In the context of chronic pain, most studies have used networks to investigate the relationships between psychological and pain variables (e.g., Åkerblom et al., 2021; Van Der Wijk et al., 2025; Zhao et al., 2023). A few studies have included one or more social and/or demographic factors next to psychological ones, and found that they are often related to pain intensity and pain disability (e.g., Chisari et al., 2021; Hinze et al., 2021). Notably, Gevers-Montero et al. (2023) observed that the factors with the strongest connections in their chronic pain network were demographic (age, gender and marital status). While these studies thus suggest that social and societal factors have unique interactions with pain outcomes next to psychological variables, the available evidence is limited.

Here, we use network analysis to explore the interplay between psychological, social and societal factors and identify how they relate to pain intensity, pain disability and quality of life. We collected self-reported data on a range of factors from a large community sample of people experiencing chronic pain (*N* = 262) through an online survey. We used cross-sectional regularized networks consisting of partial correlations to estimate unique relationships between all included variables. We first examined if a network containing only fear-avoidance and mood factors in relation to pain intensity and pain disability could replicate previously published network studies (Van Der Wijk et al., 2025; Zhao et al., 2023), and then explored what a network with factors from all measured dimensions (psychological, social and societal) could reveal. Here, we expected that social and societal variables would have unique connections with pain intensity, pain disability and quality of life beyond what psychological variables explain.

## Methods

This study was preregistered on AsPredicted (see https://aspredicted.org/8wys-csn4.pdf) prior to data collection, and updated prior to data analysis to ensure the reliability of our analyses with our achieved sample size (see https://osf.io/p3djf/?view_only=8f243e0b6eb5481087e4fb12c56f9e70).

### Participants

Participants were recruited at Maastricht University, chronic pain associations, online groups, general practitioners and physiotherapy offices in the Netherlands and Germany, and through the authors’ social networks. The survey was first assembled in English, and later translated to both Dutch and German. Participants were eligible if they were 18 years or older, spoke sufficient English, Dutch or German and had experienced persistent or recurrent pain for at least 3 months. Participants could enter a lottery with a 1/50 or higher chance to win a €50 voucher. Students from Maastricht University could choose to earn 0.5 participation credits instead. Data collection started in May 2024 and ended in August 2025.

In total, 426 participants provided written informed consent for the study. Individuals who did not meet the inclusion criteria were excluded (*N* = 8 did not experience pain in the past six months, *N* = 12 reported a pain duration of less than three months). We removed 5 duplicates. Next, we removed participants who did not fill out any of the questionnaires (*N* = 119). Additionally, we removed participants who finished the survey in less than 5 minutes (*N* = 2). None of the participants answered two or more attention checks wrong. Lastly, participants were excluded if they missed more than 50% of the items for one or more of the variables (*N* = 18). The final sample consisted of a total of *N* = 262 participants. We performed simulations prior to data collection and data analysis to ensure the expected performance of our analyses would be adequate under the study conditions (∼250 participants, 15 variables). We described these in more detail in the **Supplementary Materials**.

### Study Procedure

Participants were asked to fill in 27 questions about demographics and pain characteristics (e.g., pain duration, location, age, gender identity), and 20 questionnaires (104 items total) via Qualtrics (Provo, UT). The survey took about 30 minutes to complete. The order of the questionnaires was randomized in blocks to balance any potential impact of filling out one questionnaire before/after the other. Participants were allowed to take breaks as long as they completed the survey within one week. Participants gave informed consent before starting the survey. Ethical permission was obtained from the Ethics Review Committee Psychology and Neuroscience (ERCPN) of Maastricht University (code 281_66_04_2024).

### Questionnaires

Participants completed a series of questionnaires assessing psychological, social, and societal factors related to chronic pain. We aimed to capture a comprehensive range of aspects related to chronic pain based on the available literature. Additionally, we sought to minimize overlap between constructs and keep the assessment burden manageable for participants.

Psychological instruments were included to assess cognitive, behavioural and emotional aspects associated with chronic pain, namely pain-related worry, anxious and depressive symptoms, pain avoidance, pain-means-harm beliefs, pain acceptance, and positive affect (Alschuler et al., 2016; Droppert and Knowles, 2023; McCracken et al., 2004). Since social environments can either buffer against or exacerbate pain-related distress, influence treatment outcomes, and affect emotional well-being (Bean et al., 2022; Coady et al., 2024; Weiß et al., 2024), the social factors emotional support, experiences of discrimination and pain invalidation were assessed. Finally, to capture how structural and systemic conditions shape individuals’ experiences of pain, access to support, and overall quality of life (Atkins and Mukhida, 2022; Dorner et al., 2011; Mielck et al., 2014), measures of financial worry, stress and access to health care were included. We measured pain intensity, pain disability and quality of life as pain outcomes. The questionnaires used in this study are presented in **Table S1**.

Five attention checks were embedded throughout the survey to ensure accurate responses from participants, including three direct attention checks (e.g., *Please select “6” to show you are paying attention to this question*), one commitment question (*Do you commit to providing thoughtful answers?*), and one logic question (*Which of the following is a vegetable?*). We also included a honeypot question and CAPTCHA to prevent bots from participating.

The survey was later also administered in Dutch and German to extend data collection. Eleven questionnaires were already translated by others (see **Table S2**). For the remaining nine German and nine Dutch questionnaires, we used a forward–backward translation process, following the cross-cultural 5-step translation process by Beaton et al. (2000). This process aims to ensure that the source and target questionnaires are aligned in terms of meaning, language use and cultural relevance (Beaton et al., 2000). All translators received a small honorarium for their contribution. More information about the translation procedure can be found in the **Supplementary Materials**.

### Variables

Among the included participants, none had any missing data. For most questionnaires and subscales, the items were reverse-coded where needed and summed to calculate the value for the variable it measured. There were a few exceptions. For *experiences of discrimination*, we did not include the third item, as this was categorical. For *pain invalidation*, we combined the two subscales into a single variable by adding them up. For the Chronic Pain Grade Scale, we calculated the *Characteristic Pain Intensity* and *Disability Scores* following the instructions from the scale developers (Von Korff et al., 1992) for our *pain intensity* and *pain disability* variables, respectively. Namely, we took the average of items 1-3 and 5-7, respectively, and multiplied them by 10 to get a score between 0-100. As anxious and depressive symptoms are often strongly correlated, and to reduce the number of variables in our extended network analysis, we decided to combine the PHQ-2 and GAD-2 into a single variable reflecting *mood symptoms*, as has been done in previous studies (Kroenke et al., 2009; Pagé et al., 2021).

Most variables had good or excellent internal consistency (*Cronbach’s* α > 0.8), while *stress*, *pain intensity* and *positive affect* were acceptable (α > 0.7), and *pain acceptance, experiences of discrimination* and *pain avoidance* were somewhat low (α > 0.65) in terms of internal consistency. *Pain-means-harm beliefs* had a very low internal consistency (α = 0.12), therefore, we decided to conduct our main analyses with the single-item version of the subscale (Jensen et al., 2003). See **Table S3** for the internal consistency of all variables.

## Data Analysis

### Model assumptions

We checked for violations of normality using histograms and Q-Q plots, as well as Henze-Zirkler’s multivariate normality test from the *mvn* function in the MVN package (Korkmaz et al., 2014). None of the variables had strictly normal distributions, however, only five showed a substantial violation (skewness > ±0.5; i.e., *emotional support, depressive symptoms, pain invalidation, pain-related worry* and *experiences of discrimination*). Following recommendations based on an extensive simulation study (Isvoranu and Epskamp, 2023), we applied a non-paranormal transformation to maintain high expected model performance (precision, specificity, correlation and sensitivity to top 50% of edges > 0.8). This greatly improved data normality, and Henze-Zirkler’s test was now non-significant for both the replication (test statistic = 0.96, p = .27) and the extended (test statistic = 1.00, p = .11) networks, though *emotional support* and *experiences of discrimination* still showed substantial skewness (-0.5 and 0.4, respectively, see **Figures S1** and **S3**). We created scatter plots for each combination of variables to check for linearity, and identified no signs of violations of this assumption, apart from the ceiling and floor effects in *emotional support* and *experiences of discrimination* being visible (see **Figures S2** and **S4)**.

### Network analysis

We used Gaussian Graphical Models (GGMs), which estimate partial correlations between each combination of variables while correcting for all others. As such, the results indicate unique, direct relationships between all variables. This can be represented as a network, in which each variable is a circle (node), while each partial correlation is a line between nodes (edge; see **Figure 1**). We used the *estimateNetwork* function from the *bootnet* package in R with default settings for model estimation (Epskamp et al., 2018; Haslbeck and Waldorp, 2020). For this analysis, a series of linear multiple regressions is performed (one for each variable) in combination with least absolute shrinkage and selection operator (LASSO) regularization using 10-fold cross-validation for parameter selection. This regularization protects against issues with multicollinearity and reduces overfitting, which is especially helpful when there are many variables. Specifically, it looks for the smallest combination of predictors that can accurately predict the outcome by adding a penalty for more complex models and decreasing regression coefficients towards zero. We controlled for false positives by applying thresholding to estimates following Loh and Wainwright (Loh and Wainwright, 2012).

**Figure 1.**
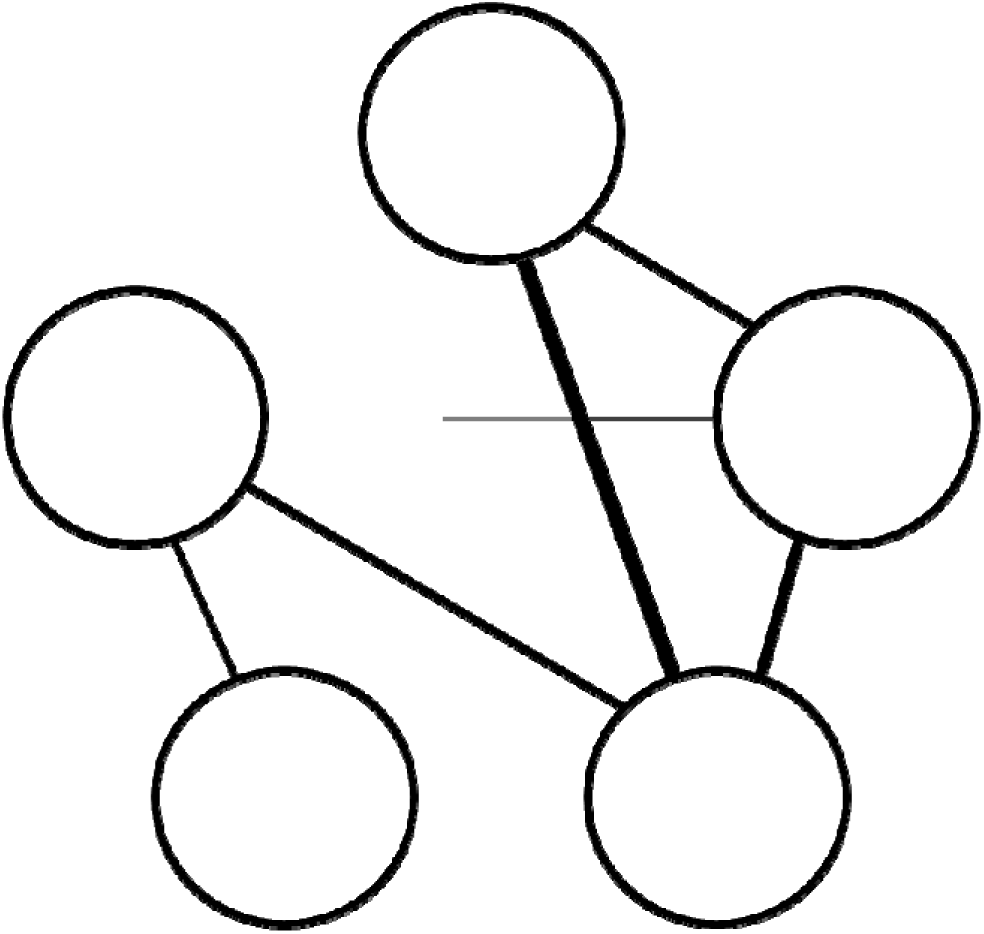
Illustration of a network model in which each variable is presented as a circle (node), while the lines between them (edges) represent the partial correlations between each pair of variables. Thicker lines indicate stronger partial correlations.

We estimated two partial correlation networks. First, we included six variables covered in two previous large-scale network studies of chronic pain (Van Der Wijk et al., 2025; Zhao et al., 2023), namely *pain intensity, pain disability, anxious symptoms, depressive symptoms, pain-means-harm beliefs* and *pain avoidance*. Then, we expanded the model to include the 15 main variables in our study: *pain intensity, pain disability, quality of life, pain-related worry, pain-means-harm beliefs, pain avoidance, pain acceptance, positive affect, mood symptoms, stress, emotional support, pain invalidation, experiences of discrimination, access to healthcare*, and *financial worry*.

### Network stability and predictability

In line with Epskamp et al. (2018), we examined the stability of the estimated networks using bootstrapping. Specifically, we repeated network estimation 5000 times with bootstrapped samples, which means that, for each sample, participants were randomly selected from the full sample with replacement. The percentage of bootstrapped samples in which an edge is identified, and the distribution of the edge estimates across bootstrapped samples provide an indication of the stability and accuracy of the edges, with higher percentages and narrower distributions pointing to more stable and accurate partial correlations. We also estimated the predictability of the nodes (Haslbeck and Fried, 2017), which represents the proportion of variance in one variable explained by all other variables (*R^2^*).

### Software and Scripts

All data cleaning and analysis was done in R Statistical Software (v4.4.1; R Core Team, 2024). The main packages we used include *mgm* (v1.2.13; Haslbeck and Waldorp, 2020), *bootne* (v1.5; Epskamp et al., 2018) and *qgraph* (v1.9.3; Epskamp et al., 2012). The scripts used in this study are available through OSF (https://osf.io/p3djf/?view_only=8f243e0b6eb5481087e4fb12c56f9e70).

### Positionality Statement

Conducting research involves making many decisions along the way. Positionality statements inform readers about the background and experiences of authors that shape the lens through which they engage with their research. While positionality statements are a longstanding practice in qualitative research (Darwin Holmes, 2020), there have been increasing calls for quantitative researchers to reflect and report on their positionality as well (Hood et al., 2022; Jamieson et al., 2022; Roberts et al., 2020). In line with these calls, we offer the following positionality statement.

We all grew up and live in Western countries and have been trained in the Eurocentric scientific tradition. While some of us have some insight into other perspectives, either through personal ties or engaging with the work of non-Western scholars, we believe the presented work largely represents Western views on science, pain and health. Most of us have a background in psychology, with one having clinical training and expertise. One author has a background in cultural anthropology, and we aimed to incorporate anthropological and sociological perspectives into this project, however, psychological perspectives provided the main guidance and direction. Our team consists of six women and one man. Five of us (all women) have lived experiences of chronic health conditions, including chronic pain. Five of us are White Europeans, one of us a Black American, and one of us comes from the Caucasian mountain area, now living in Western Europe. While we thus have some relevant experiences to health and other societal inequities, we also carry many privileges (e.g., financial stability) and do not match the diverse backgrounds and experiences of our sample.

## Results

### Sample Characteristics

The demographic and pain characteristics of our sample are presented in **Table 1**. Overall, the sample was fairly young (M = 31.1, SD = 15.2), and consisted primarily of women (78.6%) of European ethnicity (85.1%) who have (38.9%) and/or are currently working towards (studying; 52.7%) a university degree. On average, the sample had moderate pain intensity (M = 54.7, SD = 16.2) and pain disability (M = 48.6, SD = 23.8), though most of the sample had pain for over 2 years (87.2%). Participants often reported more than one pain location, most commonly in the back (45.0%) and the neck or shoulders (42.0%).

**Table 1.** Demographic and Pain Characteristics of the Sample.

| Demographic characteristics |  | Pain characteristics |  |
| --- | --- | --- | --- |
| Age* | 31.1 ± 15.2<br>(18-86) | Pain intensity | 54.7 ± 16.2<br>(10-93) |
| Biological sex |  | Pain disability | 48.6 ± 23.8<br>(0-100) |
| ☐ Female | 221 (84.7%) | Pain onset |  |
| ☐ Male | 38 (14.6%) | ☐ Gradual | 129 (49.2%) |
| ☐ Other or prefer not to answer | 3 (0.8%) | ☐ Sudden, with<br>clear reason | 50 (19.1%) |
| Gender identity |  | ☐ Sudden, without<br>clear reason | 73 (27.9%) |
| ☐ Man | 40 (15.3%) | ☐ Other | 10 (3.8%) |
| ☐ Non-binary | 10 (3.8%) | Pain duration |  |
| ☐ Woman | 206 (78.6%) | ☐ 3-6 months | 21 (8.0%) |
| ☐ Other or prefer not to answer | 6 (2.3%) | ☐ 6 months - 1 year | 16 (6.1%) |
| Racial/ethnic background* |  | ☐ 1-2 years |  |
| ☐ African | 6 (2.3%) |  |  |
| ☐ East, South and Southeast Asian | 5 (1.9%) |  |  |
| ☐ European |  |  |  |
| ☐ Latin American | 223 (85.1%) | ☐ 2-5 years | 20 (7.6%) |
| ☐ Middle Eastern and Central Asian | 1 (0.4%) | ☐ More than 5 years | 75 (28.6%) |
| ☐ Mixed European & non-European | 12 (4.6%) | Pain location | 130 (49.6%) |
| ☐ North American or Australasian | 8 (3.1%) | ☐ Arms/hands |  |
| Minority status | 6 (2.3%) | ☐ Back | 73 (27.9%) |
| ☐ No |  | ☐ Chest | 118 (45.0%) |
| ☐ Yes |  | ☐ Digestive system | 43 (16.4%) |
| Living situation* | 230 (87.8%) | ☐ Face/head | 67 (25.6%) |
| ☐ Living alone | 32 (12.2%) | ☐ Feet/legs | 103 (39.3%) |
| ☐ Living with partner and/or kids | 87 (33.2%) | ☐ Genital | 101 (38.5%) |
| ☐ Living with parents/guardians | 72 (27.5%) | ☐ Hips/knees | 22 (8.4%) |
| ☐ Living with roommates | 41 (15.6%) | ☐ Neck/shoulders | 107 (40.8%) |
| Living environment | 61 (23.3%) | ☐ Widespread | 110 (42.0%) |
| ☐ Village |  | ☐ Other |  |
| ☐ Small town |  | Health care visits | 48 (18.3%) |
| ☐ Middle-sized town | 22 (8.4%) | ☐ No | 33 (12.6%) |
| ☐ Big city | 43 (16.4%) | ☐ Yes, GP/physio-<br>therapist only | 13 (5.0%) |
| ☐ Metropolis | 74 (28.2%) | ☐ Yes, other<br>specialist(s) | 52 (19.8%) |
| Education | 99 (37.8%) | Pain diagnosis | 197 (75.2%) |
| ☐ Primary education | 24 (9.2%) | ☐ No |  |
| ☐ Secondary education |  | ☐ Yes |  |
| ☐ Post-secondary education | 2 (0.8%) | Current treatment* |  |
| ☐ Bachelor's degree or equivalent | 122 (46.6%) | ☐ No | 86 (32.8%) |
| ☐ Master's degree or equivalent | 36 (13.7%) | ☐ Yes | 176 (67.2%) |
| Employment | 56 (21.4%) | Other diagnoses |  |
| ☐ Disability benefits | 46 (17.6%) | ☐ No | 150 (57.5%) |
| ☐ Retired |  | ☐ Yes | 111 (42.5%) |
| ☐ Studying | 24 (9.2%) |  |  |
| ☐ Studying & working (paid & unpaid) | 16 (6.1%) |  |  |
| ☐ Unemployed | 105 (40.1%) |  |  |
| ☐ Unpaid work | 33 (12.6%) |  |  |
| ☐ Working full-time | 10 (3.8%) |  |  |
| ☐ Working part-time | 3 (1.1%) |  |  |
|  | 37 (14.1%) |  |  |
|  | 34 (13.0%) |  |  |

\* These variables had 1 missing value each

### Replication Network

The partial correlation network estimated from similar variables as two large sample studies (Van Der Wijk et al., 2025; Zhao et al., 2023) is presented in **Figure 2**. Seven out of the 15 possible edges were estimated to be present and all of them were positive, with partial correlations ranging from .05 (*anxious symptoms - pain intensity*) to .54 (*depressive symptoms - anxious symptoms;* see **Table S4**). The bootstrapping procedure showed that all estimated edges were present in the majority of bootstrapped samples, with the four strongest edges being present more than 97% of the time (see **Figure S5**). Furthermore, the 95% quantiles around the edges were .17-.32 wide, indicating that, in 95% of the bootstrapped samples, the estimated partial correlations fell within a ±.08-.16 window of the estimates in the main analysis of the full sample. The predictability varied from node to node, with the most variance explained for *depressive symptoms* (*R^2^* = .40 or 40%), and least amount of variance explained for *pain avoidance* (*R^2^*= .05 or 5%).

**Figure 2.**
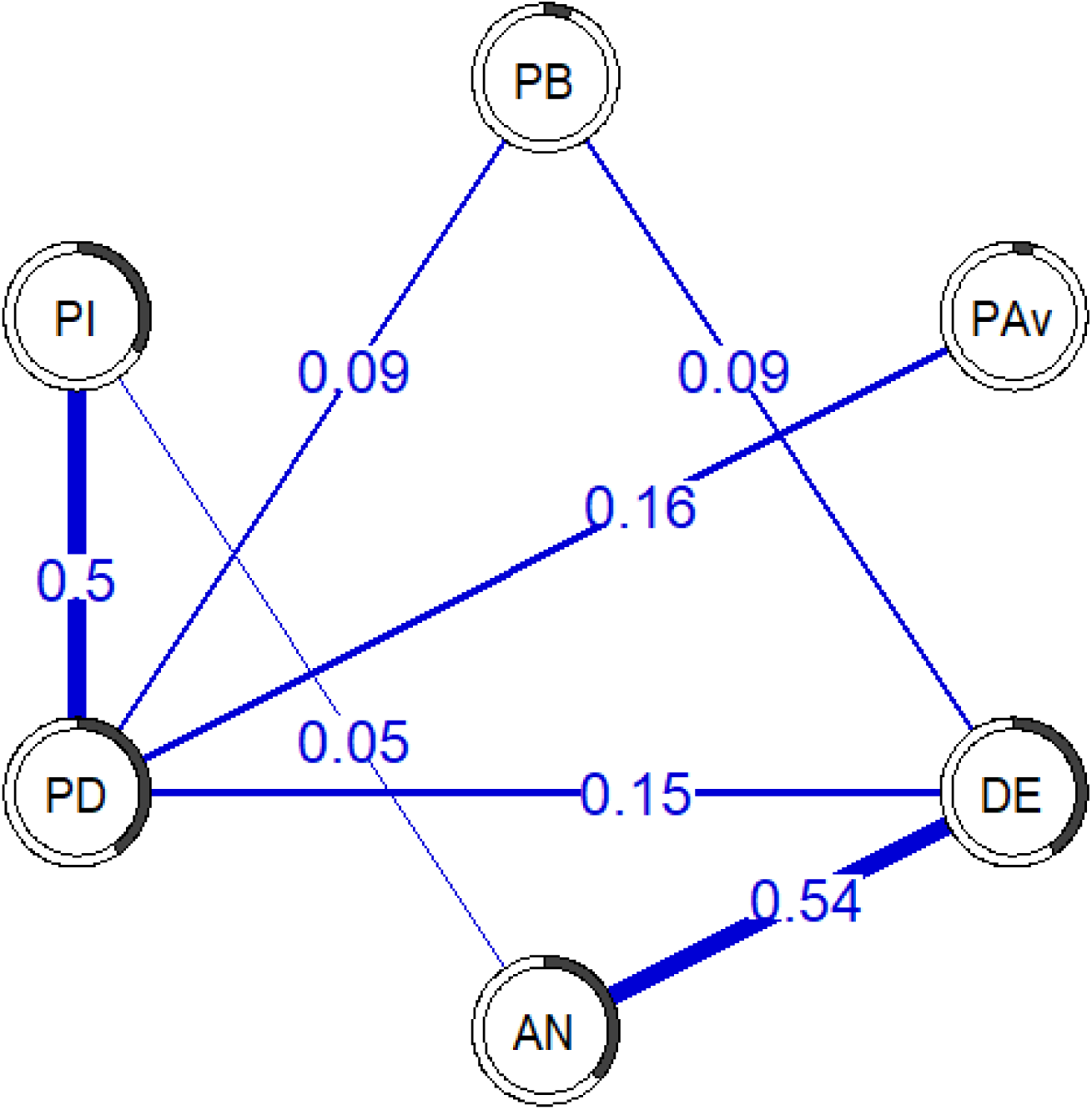
Cross-sectional replication network. The lines between variables (edges) represent partial correlations, with thicker lines indicating stronger partial correlations. The pie charts around each circle (node) represent the proportion of variance explained by the other variables (*R^2^*). PB = Pain-means-harm beliefs, PAv = Pain avoidance, DE = Depressive symptoms, AN = Anxious symptoms, PD = Pain disability, PI – Pain intensity

### Extended Network

The partial correlation network estimated from the extended set of variables is presented in **Figure 3**. Out of the 105 possible edges, 41 were estimated to be present. Twenty-four of the edges were positive, with the largest partial correlation existing between *mood symptoms* and *stress* (*r* = .46). Sixteen of the edges were negative, with the largest partial correlation existing between *pain-related worry* and *pain acceptance* (*r* = -.30). See **Table S5** for an overview of all partial correlations. Among our pain outcomes of interest, *pain intensity* had direct relationships with *pain-related worry* (*r* = .23) and *pain disability* (*r* = .35), and *pain disability* had additional direct relationships with *pain acceptance* (*r* = -.25), *pain-related worry* (*r* = .11), and *quality of life* (*r* = -.06). *Quality of life* had relationships with nine of the other variables in the network, among which *positive affect* (*r* = .30), *stress* (*r* = .-25), *financial worry* (*r* = -.23) and *experiences of discrimination* (*r* = -.20) showed the strongest associations. The bootstrapping procedure showed that all estimated edges were present in the majority of bootstrapped samples, and all edges estimated at *r* = ±.1 were present at least 75% of the time (see **Figures S6** and **S7**). In addition, the 95% quantiles around the edges were .08-.32 wide, indicating that in 95% of the bootstrapped samples, the estimated partial correlations fell within a ±.04-.16 window of the estimates in the analysis of the full sample. The predictability varied from node to node, with the most variance explained for *quality of life* (*R^2^* = .69 or 69%), and least amount of variance explained for *pain beliefs* (*R^2^*= .11 or 11%).

**Figure 3.**
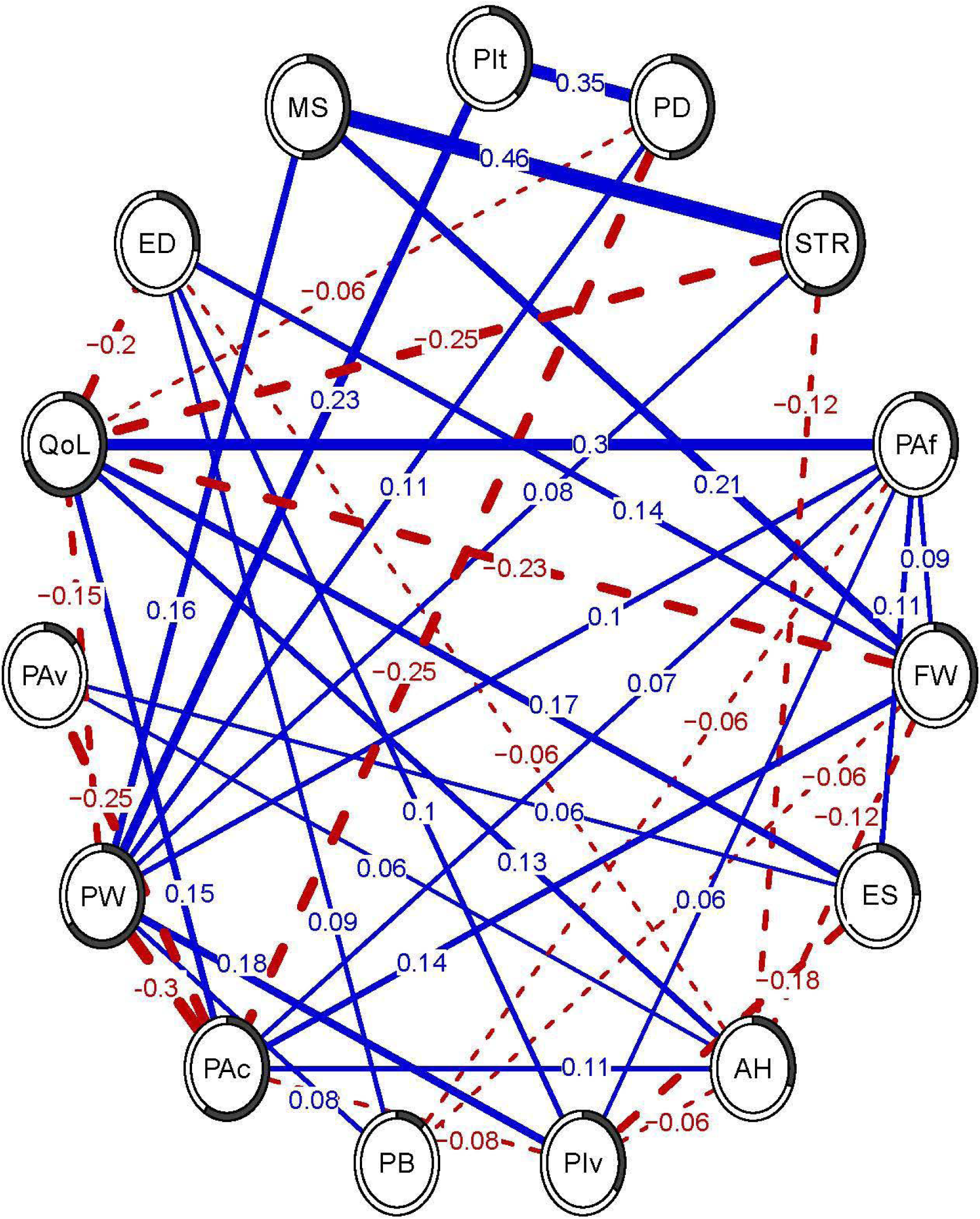
Cross-sectional extended network. The lines between variables (edges) represent partial correlations, with blue solid lines indicating positive relationships, red dashed lines indicating negative relationships, and thicker lines indicating stronger partial correlations. The pie charts around each circle (node) represent the proportion of variance explained by the other variables (*R^2^*). PIt = pain intensity, PD = pain disability, STR = stress, PAf = positive affect, FW = financial worries, ES = emotional support, AH = access to health care, PIv = pain invalidation, PB = pain-means-harm belief, Pac = pain acceptance, PW = pain-related worries, PAv = pain avoidance, QoL = quality of life, ED = experiences of discrimination, MS = mood symptoms

## Discussion

We aimed to replicate previous networks of psychological factors related to chronic pain disability, and build on this work by including social and societal factors in our extended network analysis. Despite using different measures and recruiting directly from the community instead of a specialized pain clinic, our replication network showed many similarities to previously published networks, including pain disability being related to pain intensity, depressive symptoms, pain avoidance and pain-means-harm beliefs. In our extended network, pain intensity and disability were still strongly related to each other, but now only showed direct relationships with pain-related worry and pain acceptance. Pain avoidance, pain-means-harm beliefs and depression (mood symptoms) were still indirectly related to pain disability and pain intensity, together with pain invalidation, financial worries, access to health care, stress and positive affect. Quality of life was directly related to nine of the fourteen variables, most notably positive affect, stress, financial worry and experiences of discrimination.

Our replication network was similar to two large-sample network studies using comparable variables (Van Der Wijk et al., 2025; Zhao et al., 2023), indicating a degree of stability in this pattern across samples. While our network had fewer connections overall, the strongest relationships were all replicated, with one notable exception. Namely, no relationship was identified between pain-means-harm beliefs and pain avoidance. This likely occurred because we used different measures. Notably, we measured pain avoidance through items asking how often participants avoid activities they associate with pain, whereas others (Van Der Wijk et al., 2025; Zhao et al., 2023) examined beliefs on the importance of avoiding activities (alone or in combination with the meaning of pain). Even so, both pain avoidance and pain-means-harm beliefs were related to pain disability, as in previous studies (Åkerblom et al., 2021; Thompson et al., 2019; Van Der Wijk et al., 2025; Zhao et al., 2023). Similarly, the relationships of pain disability with both pain intensity and depressive symptoms we found have commonly been observed by others as well (Åkerblom et al., 2021; Thompson et al., 2019; Van Der Wijk et al., 2025; Zhao et al., 2023). Overall, we found a similar network of psychological factors related to chronic pain, suggesting that pain disability has unique relationships with almost all of them.

In contrast, only pain acceptance and pain-related worry were directly related to pain disability in the extended network analysis. Pain avoidance, pain-means-harm beliefs and mood symptoms were related to pain acceptance and/or pain-related worry, suggesting potential indirect relationships with pain disability. A similar indirect relationship between fear-avoidance and pain disability through pain acceptance was also observed by Åckerblom et al. (2021). In addition, pain-related worry and pain acceptance were associated with pain invalidation, stress, financial worry, positive affect and access to health care, indicating that several social-societal factors may also be related to pain disability through these psychological coping strategies. In line with psychological research on chronic pain, our analysis highlighted core concepts from two distinct psychological models of chronic pain (i.e., the psychological flexibility model and fear-avoidance model; McCracken and Morley, 2014; Vlaeyen and Linton, 2012) as being the most directly related to pain disability and pain intensity. Overall, these results suggests that pain acceptance and pain-related worry play a primary role in understanding pain disability, while other psychological and social-societal factors play an indirect role.

This pattern invites us to look beyond the immediate psychological factors to what may shape individual differences in these factors. For example, our results indicate that invalidating responses from others are associated with more pain-related worry and less pain acceptance. Although our cross-sectional design leaves the direction of influence open, several qualitative studies have highlighted a lack of understanding and support from others as a barrier to pain acceptance (Biguet et al., 2016; LaChapelle et al., 2008), suggesting a causal effect. While we know of no studies examining the relationship of pain acceptance with stress, access to health care and financial worry, theoretical models of pain and health inequities have proposed that lower socio-economic position exerts its negative effect through shaping coping strategies (Kapos et al., 2024; Khalatbari-Soltani and Blyth, 2022; Taylor et al., 1997). Our findings align with this idea that responses to chronic pain, such as worry and acceptance, are impacted by the social context and broader societal structures people live in.

The importance of social and societal factors is further emphasized by the substantial direct relationships of quality of life with stress, financial worries, experiences of discrimination, emotional support and access to health care. Notably, these factors seemed to have stronger relationships with quality of life than pain disability and pain intensity. Furthermore, these relationships appeared despite having a relatively well-resourced and connected sample (e.g., high social support, low financial worry). We would expect the strength of such relationships to be even more pronounced in more diverse samples.

Since quality of life is an important outcome of treatment (e.g., Cohen et al., 2021; Scascighini et al., 2008), our findings indicate that taking social and societal factors into account during treatment alongside more typically targeted psychological factors could lead to more effective interventions, as others have suggested (Kapos et al., 2024; Slater et al., 2024; Wallace et al., 2021). Although societal factors are more difficult to address in individual interventions, several models have been developed to highlight health care practices that may reduce the negative impacts of societal stressors for patients undergoing treatment (e.g., RESTORATIVE model, Hood et al., 2023; EQUIP Healthcare model, EQUIP Research Team et al., 2015). A study implementing the EQUIP Healthcare model in four primary care clinics found that higher levels of equity-oriented care was associated with better health outcomes (FORD-GILBOE et al., 2018). Interestingly, this association was mediated by patient confidence in managing their health, highlighting a potential psychological mechanism, similar to our findings.

Addressing the root causes of such social-societal impacts would likely be even more effective, as others have put forward as well (e.g., Potthoff et al., 2025). For example, Hood et al. (2023) emphasize that the most impactful actions for addressing race-related pain inequities require structural changes from health care providers and the institutions they work at to prevent racialized people from being exposed to discrimination. Longitudinal findings that more financial worry was associated with higher pain ratings on a daily level, suggest that alleviating economic hardship could also reduce chronic pain (Rios and Zautra, 2011). While this is challenging, long-term work, several initiatives have already been taken to start addressing societal causes of pain inequities. For example, Hirsch et al. (2019) developed and tested a virtual intervention to reduce treatment bias related to race and socio-economic position in health care providers. Though not specific to chronic pain, the need for policy and political interventions to address health inequities has been emphasized, for example by improving access to affordable care, housing and education (Baum, 2016).

More work on the interactions between social-societal and psychological factors is needed to illuminate the direction and nuances of their interactions. Our study provides preliminary insights by highlighting how specific social-societal and psychological factors interact, pointing to potential mechanisms. Since our survey method provided only surface level information on a single point in time, much remains unknown about the direction and meaning of our findings. For example, we were surprised that more financial worry was associated with more pain acceptance, but our online survey method does not allow for in-depth understanding. Seeing the complex and multifactorial nature of chronic pain, we agree with previous work (Hood et al., 2023; Kapos et al., 2024) that interdisciplinary collaborations across the fields of public health, medical anthropology, psychology and sociology could provide the insights needed to guide future efforts.

Our study has several limitations. While both a-priori simulations and post-hoc stability measures indicated our sample size was adequate, the accuracy of our network estimates was somewhat lower than in previous studies with larger samples. We managed to recruit a community sample diverse in their pain characteristics; however, it was considerably skewed in sociodemographic characteristics, most notably consisting largely of young white women with or on track to have university-level education. In addition, our recruitment and data collection methods may have encouraged participation from those most interested in sharing their pain experiences, and may have discouraged those with fewer digital skills and/or who do not speak English, Dutch or German. As such, our results may not generalize to the whole population of people experiencing chronic pain. Our use of a one-time online survey further limited our ability to make causal inferences. We selected short-form questionnaires where possible to reduce the participant burden, however, these may have provided a limited estimate of the factors of interest. This is especially likely for the pain-means-harm variable, as we ended up using the 1-item version due to low internal consistency for the 2-item version in our sample. Even so, the survey was relatively long, which may have led to motivation and attention dropping towards the end. Lastly, the items for pain avoidance were accidentally measured on a 4-point instead of a 5-point scale, deviating from the original Activity Patterns Scale (Esteve et al., 2016).

In conclusion, our results emphasize that, when taking important psychological factors into account, social and societal factors remain related to important pain outcomes. Specifically, our results showed that psychological, social and societal factors all directly relate to quality of life among people with chronic pain, with stress, financial worry, positive affect and experiences of discrimination being the most prominent. Pain disability and pain intensity only had direct relationships with pain-related worry and pain acceptance, however, these factors were in turn related to social and societal factors, suggesting that the context people live in can act as a barrier or facilitator for better pain management. Future work should examine the interactions between psychological and social-societal factors in more depth to understand how they shape pain coping responses and can thus be best approached through intervention. In addition, our findings emphasize the need for structural changes (e.g., addressing discrimination, pain invalidation and socio-economic inequities) to alleviate the burden of chronic pain.

## Acknowledgements

We would like to thank Verena Hofmann, Anne Lukas, Hannah Marten, Malcom Sepulchre, Niels Weber, Yari Msika, Mikal Weldeyesus, Thomas Vermeulen, Johan Vlaeyen and Madelon Peters for their advice in setting up the survey and/or help with translating the survey faithfully. This study was preregistered on OSF (https://osf.io/p3djf/?view_only=8f243e0b6eb5481087e4fb12c56f9e70).

## Data Availability

The data used in this study is available on OSF: https://osf.io/p3djf/?view_only=8f243e0b6eb5481087e4fb12c56f9e70.

