## Supplementary Materials for "Social and Societal Factors Interact with Psychological Factors to Shape Pain Outcomes in a Community Sample with Chronic Pain: A Network Study"

1. **Simulations**

We performed simulations to estimate the performance of network analyses depending on sample size (100, 250, 350, 500, 750, or 1000 participants), number of variables (5, 10, 15, 20, or 25) and network characteristics (low, medium, or high density). More specifically, we constructed networks for each combination of conditions to serve as ‘true’ networks. We then simulated data based on these network parameters, used network analyses to estimate networks from the simulated data, and compared the estimated networks to the ‘true’ networks. We used four measures of performance, the estimation error (mean absolute difference between estimated and ‘true’ networks), sensitivity (true positives), specificity (true negatives) and the correlation between the simulated and ‘true’ network. The simulations were repeated 100 times for each combination of different sample sizes, numbers of variables and network characteristics. The script we used to perform these simulations can be found on OSF (<https://osf.io/p3djf/overview?view_only=8f243e0b6eb5481087e4fb12c56f9e70>).

While we originally aimed to collect data from 500 participants, providing adequate performance for networks with 25 variables (see <https://aspredicted.org/8wys-csn4.pdf>), we were only able to include ~250 participants. Therefore, we reduced the number of variables included in the main analysis to 15. Averaging over repetitions, the simulations indicated that this should provide great performance for low density networks (error = 0.01, sensitivity = 0.99. specificity = 0.96, correlation = 0.98), good performance for medium density networks (error = 0.02, sensitivity = 0.88, specificity = 0.89, correlation = 0.90) and decent performance for high density networks (error = 0.04, sensitivity = 0.42, specificity = 0.85, correlation = 0.52).

2. **Questionnaires**

**Table S1**

*Summary of Study Variables, Questionnaires, and their Psychometric Properties*

| **Variable** | **Questionnaire** | **No of Items** | **Answer scale** | **Validity and reliability** | **References** |
| --- | --- | --- | --- | --- | --- |
| *Questionnaires used in extended network analysis* | | | | | |
| Access to Health Care | Subscales Accessibility and Convenience of the Patient Satisfaction Questionnaire Short Form (PSQ-18) | 4 | 1 (strongly  agree) to 5 (strongly disagree) | α = .75  adequate construct validity | (Marshall and Hays, 1994) |
| Anxious Symptoms | General Anxiety Disorder 2-item scale  (GAD-2) | 2 | 0 (not at all) to 3 (nearly everyday) | α = .69  r =.69  good construct validity | (Hitchon et al., 2020; Plummer et al., 2016) |
| Coping Strategies | Subscales pain avoidance, excessive persistence, pacing to conserve energy for valued activities of the Activity Patterns Scale (APS) | 9 (3 per subscale)^a^ | 0 (not at all) to 4 (always)^b^ | α = .67-.91  r = .72-.82  good construct validity | (Esteve et al., 2016; Hotz-Boendermaker et al., 2024) |
| Depressive Symptoms | Patient-Health Questionnaire-2 (PHQ-2) | 2 | 0 (not at all) to 3 (nearly everyday) | α = 0.76–.83  r = .70–.92  good convergent, construct and criterion validity | (Gelaye et al., 2016; Kroenke et al., 2003; Löwe et al., 2005; Maroufizadeh et al., 2019; Yu et al., 2011) |
| Emotional Support | Patient-Reported Outcomes Measurement Information System (PROMIS) Emotional Support – Short Form 4a, v2.0 for adults | 4 | 1 (never) to 5 (always) | α = .90–.93  good criterion and construct validity | (Cella et al., 2007; Hahn et al., 2014) |
| Experiences of Discrimination | Everyday Discrimination Scale short form | 3 | First 2 items:  1 (almost everyday) to 6 (never)  Third item: asks about main reasons for these experiences, if relevant. | α = .77  r=.69 | (Benner et al., 2024; Krieger et al., 2005) |
| Financial Worry | Financial Anxiety Scale (FAS-2) | 2 | 1 (never) to 7 (always) | α = .94  good construct validity | (Archuleta, K.L. et al., 2013) |
| Pain Acceptance | Chronic Pain Acceptance Questionnaires (CPAQ-2) | 2 | 0 (never true) to 6 (always true) | accounted for 61% of variance in the 20-item scale (p < .001) | (Vowles et al., 2020) |
| Pain-Means-Harm Beliefs | 2-item version of the harm subscale of the Survey of Pain Attitudes (SOPA) | 2 | 0 (very untrue) to 4 (very true) | strong association (r = .81-.83) with the long form of the subscale  original SOPA:  α = .71 - .81  r= .63 - .68 | (Jensen et al., 2003) |
| Pain Intensity and Disability | Chronic Pain Grade Scale  (CPGS) | 7 (3 items for pain intensity, 4 items for pain disability) | Pain Intensity: 0 (no pain) to 10 (pain as bad as could be)  Disability: 0 (e.g., no interference/ no change) to 10 (e.g., extreme interference/ extreme change) | α=.67-.9132  high criterion and construct validity (including good convergent and discriminant validity) | (Smith et al., 1997; Von Korff et al., 1992) |
| Pain Invalidation | Subscales invalidation by immediate others and invalidation by health care providers of the Pain Invalidation Scale | 12 | 1 (strongly disagree) to 7 (strongly agree) | By immediate others:  α =.92, r =.75  By health care providers:  α =.93,r =.69 | (Nicola et al., 2022) |
| Pain-related Worry | Concerns About Pain (CAP) Scale | 6 | 1 (never) to 5 (always) | r=.70  strong construct validity | (Amtmann et al., 2018) |
| Stress | Perceived Stress Scale (4 item version) | 4 | 0 (never) to 4 (very often) | α =.74  Spearman-Brown split-half reliability = .76 | (Vallejo et al., 2018) |
| Positive Affect | Positive affect subscale of the short form of the Positive and Negative Affect Schedule (PANAS) | 5 | 1 (never) to 5 (always) | α=.78  r =.84 | (Thompson, 2007) |
| Quality of Life | EUROHIS-QOL | 8 | 1 (not at all) to 5 (completely) | α = .83  good structural, convergent, and discriminant validity | (Schmidt et al., 2006) |
| *Questionnaires excluded from extended analysis* | | | | | |
| Internalised Stigma | Alienation subscale of Internalised Stigma in Chronic Pain Scale | 6 | 1 (strongly disagree) to 4 (strongly agree) | α=.90 | Waugh et al. (2014) |
| Optimism | Positively worded items of the Revised Life Orientation Test (LOT-R)* | 3 | 0 (strongly disagree) to 4 (strongly agree) | α = .71  r = .79 | (Herzberg et al., 2006; Scheier et al., 1994) |
| Pain Resilience | Pain Resilience Scale (PRS) | 12 | 0 (not at all) to 4 (all the time) | α = .93  r = .80  acceptable convergent and external validity | (Ankawi et al., 2017; Slepian et al., 2016) |
| Patient-Doctor Relationship | Patient-Doctor Relationship Questionnaire (PDRQ-9) | 9 | 1 (not at all appropriate to 5 (totally appropriate) | α = .94-.96  r = .61  good convergent and discriminant validity | (Porcerelli et al., 2014; Van der Feltz-Cornelis et al., 2004; Zenger et al., 2014) |
| Trauma | Adverse Childhood Experiences Questionnaire for Adults – Short Form (ACE-Q-2) | 2 | Checkboxes (either applies or not) | α = .76–.83^c^  r = .71–.913^c^ | (Irshad and Lone, 2025; Schauss et al., 2021; Wade et al., 2017; Zanotti et al., 2018) |
| Belief in a Just World^d^ | The personal belief in a just world scale, justice subscale (PJW-O) | 8 | 1 (strongly disagree) to 6 (strongly  agree). | α = .68,  r = .57 | (Bègue and Bastounis, 2003; Dalbert, 1999) |

*Note.* This table presents the internal consistency (α) and test–retest reliability (r) of the questionnaires in previous studies.

*^a^* *Only the three items from the pain avoidance subscale were used in the main analyses*

*^b^ By accident, the current questionnaire was set up with 4 instead of 5 answer options for this scale*

*^c^ Based on ACE-10 reliability data, as data for the ACE-Q-2 are limited*

*^d^ This questionnaire was excluded from the survey when we added the German and Dutch translations, as the translation process revealed that the items from this questionnaire were difficult to translate and confusing for participants.*

### 3. **Dutch and German survey**

The English survey was later administered in Dutch and German to extend data collection to the local Dutch- and German population. Allowing participants to fill out the questionnaire in their native language improves data quality, since this lowers the probability of straight-lining or choosing default answer options (Wenz et al., 2021). Sociodemographic and pain-related clinical questions were translated by two co-authors of this study. For the questionnaires, we used already translated Dutch and German versions of the scales included in this survey where possible (see **Table S2**). For the remaining questionnaires, we followed the cross-cultural 5-step translation process by Beaton et al. (Beaton et al., 2000) to perform our own translation.

In the first step, two native Dutch and two native German speakers fluent in English independently translated the questionnaires. Two of the authors acted as forward translators, one in each language. In step 2, the translators compared their versions and discussed discrepancies until they reached a consensus on a single version per language. Next, two native English-speaking back-translators that were proficient in Dutch or German, translated the items back to English. Discrepancies between the backtranslations as well as the backtranslations and original items were documented. All translators and an additional two chronic pain experts per language evaluated the translations for accuracy, clarity and fidelity through an online survey in Qualtrics (Provo, UT). In the fourth step, all translators and chronic pain experts were invited to a meeting (one per language) to discuss the items with the most divergent quality ratings or biggest discrepancies in the backtranslations. The original translations were adjusted based on these discussions. Since we used data collected in multiple languages in one analysis, we also evaluated the existing translations and made a few minor adjustments to ensure the English, Dutch and German translations were as similar as possible. In the last step, these pre-final versions were piloted with four individuals per language who experience chronic pain to evaluate clarity, relevance, and comprehensibility. Their feedback was incorporated to improve the final versions, ensuring cultural appropriateness and content validity (Beaton et al., 2000). The final translations can be requested from the corresponding author. In the study sample, 70.2% (*N* = 184) participants filled the survey out in English 21.0% (*N* = 55) in German, and 8.8% (*N* = 23) participants in Dutch.

### 4. **Available translations**

**Table S2**

*Overview of Available Translations Used in this Study*

| **Questionnaire** | **German version** | **Dutch version** |
| --- | --- | --- |
| Chronic Pain Grade Scale | (Klasen et al., 2004) | NA – translated by authors |
| Perceived Stress Scale - 4 | Translation by Jan Engling, from [https://www.cmu.edu/dietrich/ psychology/stress-immunity-disease-lab/scales/index.html](https://www.cmu.edu/dietrich/%20psychology/stress-immunity-disease-lab/scales/index.html) | Longitudinal Aging Study Amsterdam [https://lasa-vu.nl/wp-content/ uploads/2021/03/LASA104_quest_nl.pdf](https://lasa-vu.nl/wp-content/uploads/2021/03/LASA104_quest_nl.pdf) |
| Revised Life Orientation Test (LOT-R) | (Glaesmer et al., 2008) | (Klooster et al., 2010) |
| Positive and Negative Affect Schedule (PANAS) – short form | (Röcke and Grühn, 2003) | (Engelen, Ute et al., n.d.) |
| Patient-Health Questionnaire-2 (PHQ-2) | (Löwe et al., 2010) | <https://www.phqscreeners.com/> |
| Adverse Childhood Experiences Questionnaire for Adults – Short Form (ACE-Q-2) | (Wingenfeld et al., 2010) | NA – translated by authors |
| General Anxiety Disorder 2-item scale (GAD-2) | (Löwe et al., 2008) | ECFS Mental Health Working Group [https://www.ecfs.eu/sites/default/files/ general-content-files/working-groups/Mental%20Health/GAD7 _Dutch%20for%20Belgium.pdf](https://www.ecfs.eu/sites/default/files/general-content-files/working-groups/Mental%20Health/GAD7_Dutch%20for%20Belgium.pdf) |
| Patient-Doctor Relationship Questionnaire (PDRQ-9) | (Zenger et al., 2014) | (Van der Feltz-Cornelis et al., 2004) |
| Chronic Pain Acceptance Questionnaires (CPAQ-2) | (Nilges et al., 2007) | (Trompetter et al., 2011) |
| Activity Patterns Scale | (Hotz-Boendermaker et al., 2024) | Received from authors of (Esteve et al., 2016) |
| EUROHIS-QOL | (Brähler et al., 2007) | (De Vries and Van Heck, 1996) |
| Patient-Reported Outcomes Measurement Information System (PROMIS) Emotional Support – Short Form 4a, v2.0 for adults, Dutch-Flamish | NA – translated by authors | (Haverman et al., 2016) |
| Patient Satisfaction Questionnaire | NA – translated by authors | Quakunde ([https://www.quantitativeskills.com/ quakunde/PSQ18NL.pdf](https://www.quantitativeskills.com/quakunde/PSQ18NL.pdf))  Original from RAND Health (<https://www.rand.org/health/surveys/psq.html>) |

5. **Internal consistency**

**Table S3**

*Internal Consistency of Variables Used in Analyses*

| **Variable** | **Cronbach’s alpha** |
| --- | --- |
| *Access to Health Care* | 0.80 |
| *Anxious symptoms* | 0.84 |
| *Depressive symptoms* | 0.81 |
| *Emotional Support* | 0.94 |
| *Experiences of Discrimination* | 0.69 |
| *Financial Worry* | 0.89 |
| *Pain Acceptance* | 0.67 |
| *Pain Avoidance* | 0.67 |
| *Pain Beliefs* | 0.12 |
| *Pain Disability* | 0.88 |
| *Pain Intensity* | 0.73 |
| *Pain Invalidation* | 0.92 |
| *Pain-related worry* | 0.89 |
| *Mood symptoms* | 0.84 |
| *Stress* | 0.79 |
| *Positive Affect* | 0.76 |
| *Quality of Life* | 0.83 |

6. **Model assumptions**

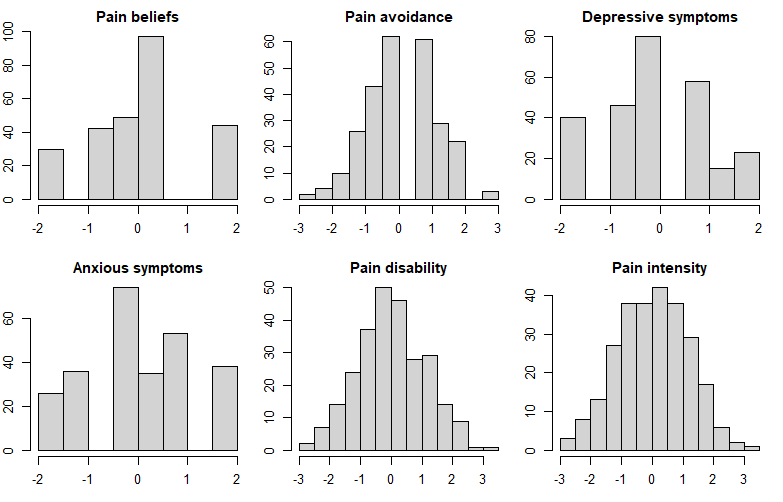

**Figure S1.** Histograms of variables used in replication network analysis after non-paranormal transformation.

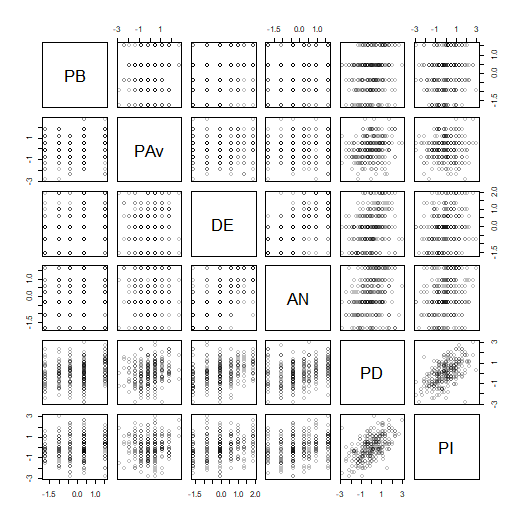

**Figure S2.** Pair-wise scatter plots of variables used in replication network analysis after non-paranormal transformation. PB = Pain-means-harm beliefs, PAv = Pain avoidance, DE = Depressive symptoms, AN = Anxious symptoms, PD = Pain disability, PI – Pain intensity

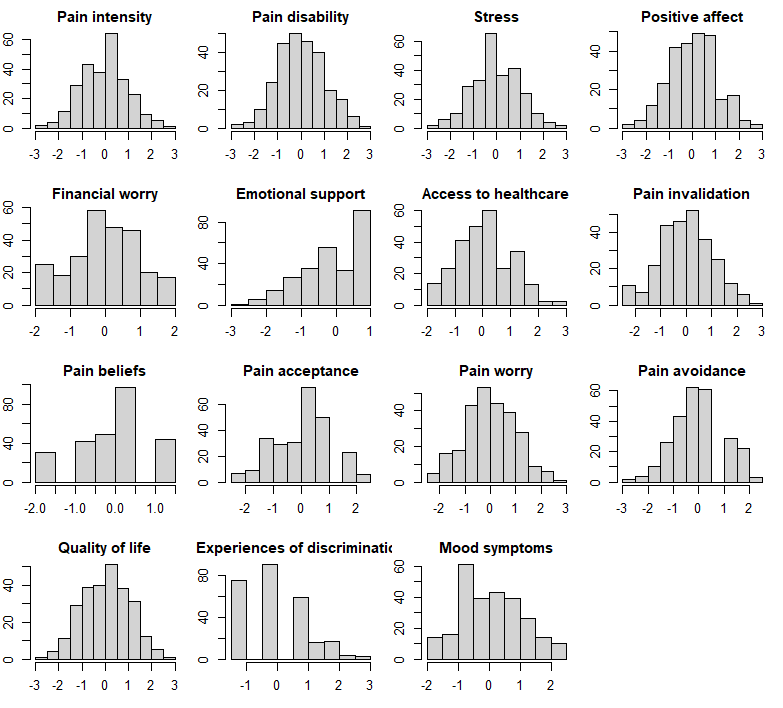

**Figure S3.** Histograms of variables used in extended network analysis after non-paranormal transformation.

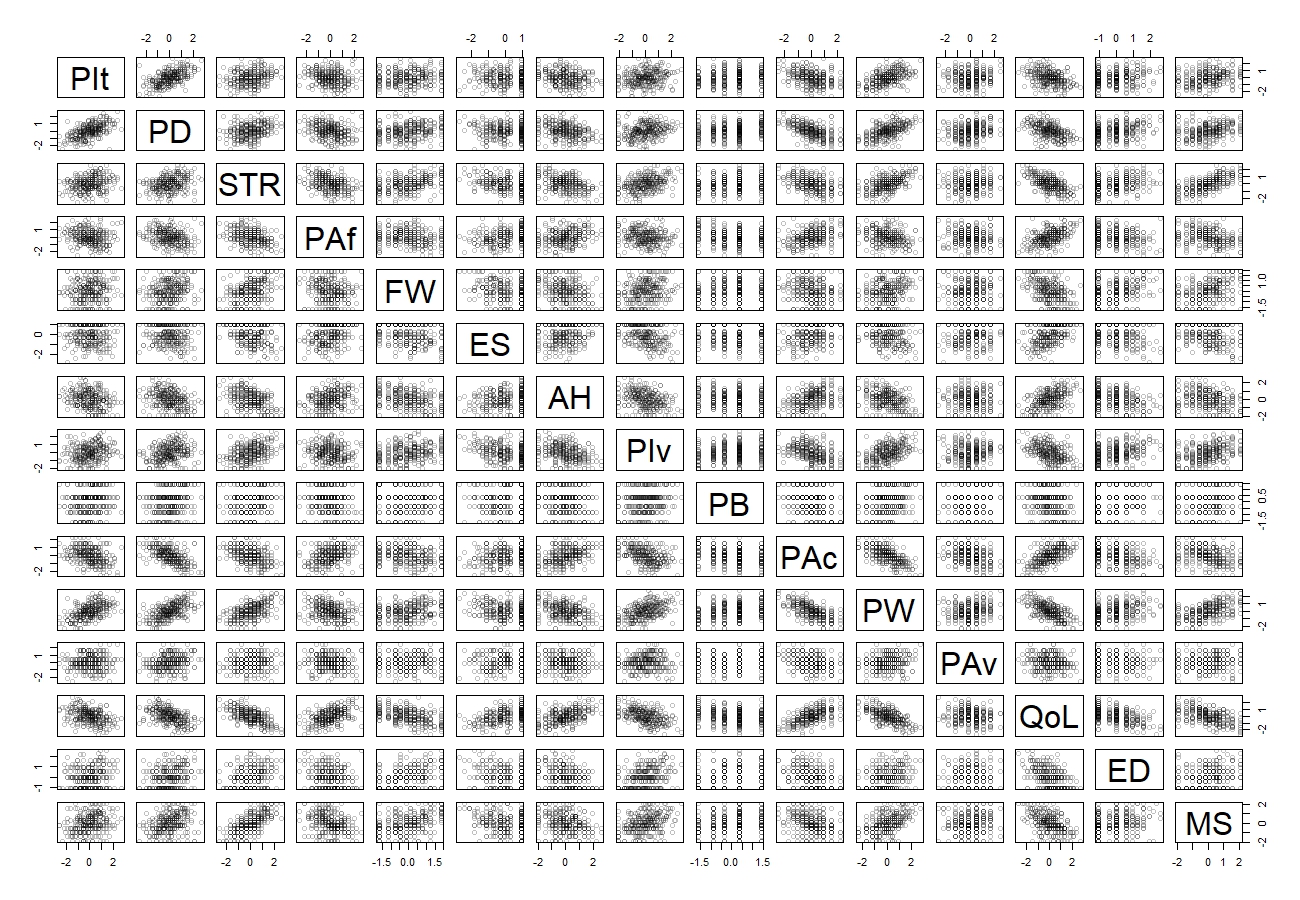

**Figure S4.** Pair-wise scatter plots of variables used in extended network analysis after non-paranormal transformation. PIt = pain intensity, PD = pain disability, STR = stress, PAf = positive affect, FW = financial worries, ES = emotional support, AH = access to health care, PIv = pain invalidation, PB = pain-means-harm belief, PAc = pain acceptance, PW = pain-related worries, PAv = pain avoidance, QoL = quality of life, ED = experiences of discrimination, MS = mood symptoms

7. **Partial correlation tables for replication and extended networks**

**Table S4**

*Partial Correlations in Replication Network*

|  | **PB** | **PAv** | **DE** | **AN** | **PD** | **PI** |
| --- | --- | --- | --- | --- | --- | --- |
| **PB** | . | . | .09 | . | .09 | . |
| **PAv** | . | . | . | . | .16 | . |
| **DE** | .09 | . | . | .54 | .15 | . |
| **AN** | . | . | .54 | . | . | .05 |
| **PD** | .09 | .16 | .15 | . | . | .5 |
| **PI** | . | . | . | .05 | .5 | . |

*Note.* PB = Pain-means-harm belief, PAv = Pain Avoidance, DE = Depressive symptoms, AN = Anxious symptoms, PD = Pain Disability, PI – Pain Intensity

**Table S5**

*Partial Correlations in Extended Network*

|  | **PIt** | **PD** | **STR** | **PAf** | **FW** | **ES** | **AH** | **PIv** | **PB** | **PAc** | **PW** | **PAv** | **QoL** | **ED** | **MS** |
| --- | --- | --- | --- | --- | --- | --- | --- | --- | --- | --- | --- | --- | --- | --- | --- |
| **PIt** | . | .35 | . | . | . | . | . | . | . | . | .23 | . | . | . | . |
| **PD** | .35 | . | . | . | . | . | . | . | . | -.25 | .11 | . | -.06 | . | . |
| **STR** | . | . | . | . | . | . | -.12 | . | . | . | .08 | . | -.25 | . | .46 |
| **PAf** | . | . | . | . | .09 | .11 | . | .06 | -.06 | .07 | .1 | . | .3 | . | . |
| **FW** | . | . | . | .09 | . | . | -.12 | . | -.06 | .14 | . | . | -.23 | .14 | .21 |
| **ES** | . | . | . | .11 | . | . | . | -.18 | . | . | . | .06 | .17 | . | . |
| **AH** | . | . | -.12 | . | -.12 | . | . | -.06 | . | .11 | . | .06 | .13 | -.06 | . |
| **PIv** | . | . | . | .06 | . | -.18 | -.06 | . | . | -.08 | .18 | . | . | .1 | . |
| **PB** | . | . | . | -.06 | -.06 | . | . | . | . | . | .08 | . | . | .09 | . |
| **PAc** | . | -.25 | . | .07 | .14 | . | .11 | -.08 | . | . | -.3 | -.25 | .15 | . | . |
| **PW** | .23 | .11 | .08 | .1 | . | . | . | .18 | .08 | -.3 | . | . | -.15 | . | .16 |
| **PAv** | . | . | . | . | . | .06 | .06 | . | . | -.25 | . | . | . | . | . |
| **QoL** | . | -.06 | -.25 | .3 | -.23 | .17 | .13 | . | . | .15 | -.15 | . | . | -.2 | . |
| **ED** | . | . | . | . | .14 | . | -.06 | .1 | .09 | . | . | . | -.2 | . | . |
| **MS** | . | . | .46 | . | .21 | . | . | . | . | . | .16 | . | . | . | . |

*Note.* PIt = pain intensity, PD = pain disability, STR = stress, PAf = positive affect, FW = financial worries, ES = emotional support, AH = access to health care, PIv = pain invalidation, PB = pain-means-harm belief, PAc = pain acceptance, PW = pain-related worries, PAv = pain avoidance, QoL = quality of life, ED = experiences of discrimination, MS = mood symptoms

8. **Post-hoc network accuracy & stability**

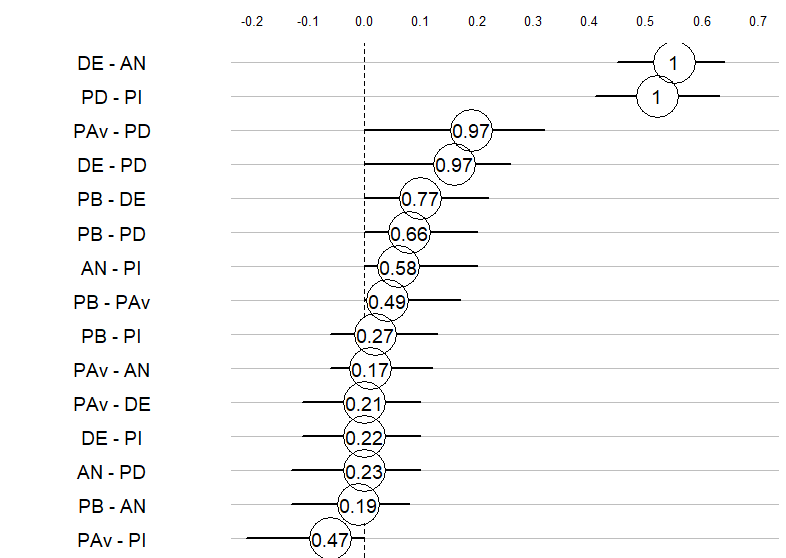

**Figure S5.** Results from the bootstrapping procedure for the replication network. Each line represents the results for one edge (estimated partial correlation). The location of the circle indicates the average estimate for that edge, while the solid line extending from the circle indicates the 95% quantiles around the estimate. The number inside the circle indicates the proportion of bootstrapped samples in which the edge was estimated to be present (e.g., 1 = 100% of the time). PB = Pain-means-harm beliefs, PAv = Pain avoidance, DE = Depressive symptoms, AN = Anxious symptoms, PD = Pain disability, PI – Pain intensity

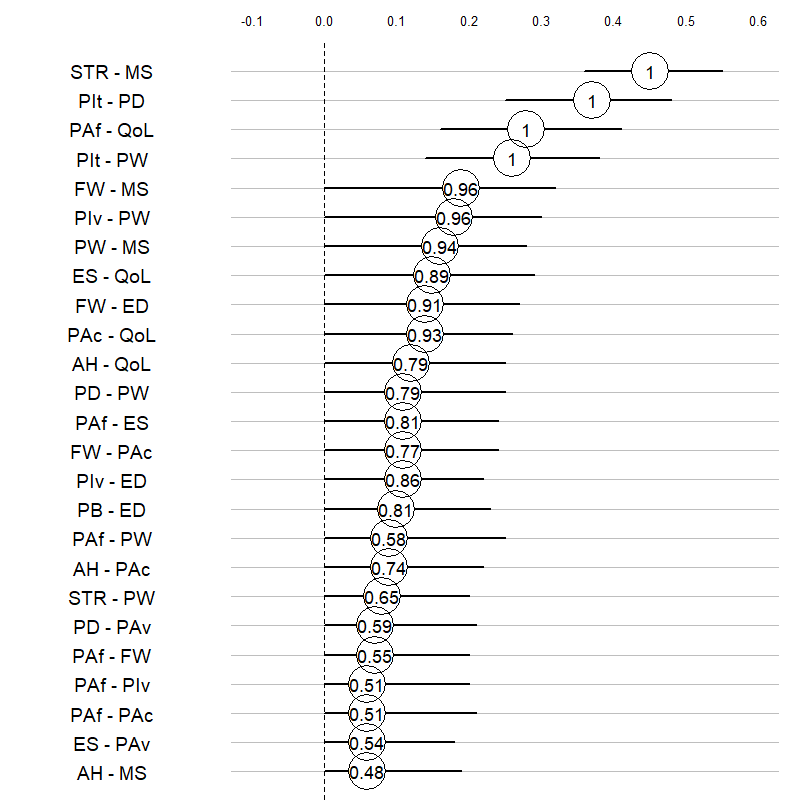

**Figure S6.** Results from the bootstrapping procedure for the 25 largest positive edges in the extended network, including the 24 edges that were estimated to be present in the main analysis. Each line represents the results for one edge (estimated partial correlation). The location of the circle indicates the average estimate for that edge, while the solid line extending from the circle indicates the 95% quantiles around the estimate. The number inside the circle indicates the proportion of bootstrapped samples in which the edge was estimated to be present (e.g., 1 = 100% of the time).

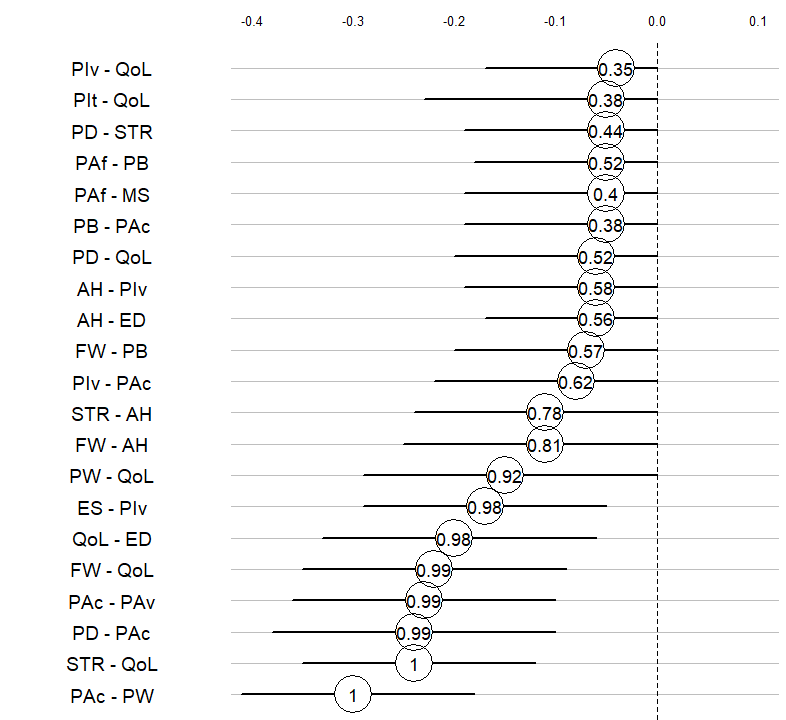

**Figure S7.** Results from the bootstrapping procedure for the 20 largest negative edges in the extended network, including the 16 edges that were estimated to be present in the main analysis. Each line represents the results for one edge (estimated partial correlation). The location of the circle indicates the average estimate for that edge, while the solid line extending from the circle indicates the 95% quantiles around the estimate. The number inside the circle indicates the proportion of bootstrapped samples in which the edge was estimated to be present (e.g., 1 = 100% of the time).
